# R21/Matrix-M Protects Against Dermal but not Against Venous Parasites in Human Challenge Studies

**DOI:** 10.1101/2024.08.06.24311495

**Authors:** Melissa C Kapulu, Francesca Orenge, Domtila Kimani, Elizabeth Kibwana, Hillary Kibet, Mary Mutahi, Mehreen S Datoo, Duncan Bellamy, Janet Musembi, Omar Ngoto, Hamisi Rashid, Stellamaris Akinyi, Mwaganyuma H Mwatasa, Lydia Nyamako, Kelvias Keter, Rose Gatheru, Agnes Mutiso, Jennifer Musyoki, Jedidah Mwacharo, Yonas Abebe, Eric J James, Peter F Billingsley, Caroline Ngetsa, Moses Mosobo, Johnstone Makale, Brian Tawa, Kevin Wamae, Lynette I Ochola-Oyier, Alison Lawrie, Fernando Ramos-Lopez, Rachel Roberts, Thomas L Richie, B. Kim Lee Sim, Stephen L Hoffman, Katie J Ewer, Adrian V S Hill, Mainga Hamaluba, Philip Bejon

## Abstract

Both licensed *Plasmodium falciparum* malaria vaccines induce anti-circumsporozoite protein antibodies that block the malaria sporozoites injected by mosquitoes. Animal models show that sporozoites in the skin are more readily blocked than intravenous sporozoites.

Controlled human malaria infections (CHMI) is usually done using infectious mosquito bites to predict efficacy in the field, but mosquito bites deliver a mixture of sporozoites into dermal layers and into capillaries.

We undertook CHMI in volunteers vaccinated with the CSP-based R21/Matrix-M vaccine or with ME-TRAP-based viral vectored vaccines. R21/Matrix-M was highly protective against CHMI using intradermal inoculation of sporozoites (i.e. 0 out of 12 volunteers met treatment criteria) but not protective against direct venous inoculation (i.e. 5 out of 5 volunteers met treatment criteria). Volunteers vaccinated with viral vectors encoding ME-TRAP antigens were not protected against intradermal inoculation.

Defining protective efficacy separately against direct venous inoculation and against intradermal inoculation enhances our mechanistic understanding of vaccination.

The study was registered with the ClinicalTrials.gov (NCT03947190) and with PACTR (PACTR202108505632810).

## Introduction

Falciparum malaria remains a pressing public health problem. It is hoped that the recently stalled progress in malaria control might be renewed by the recent approval of two efficacious vaccines (i.e. RTS,S/AS01 and R21/Matrix-M). Both vaccines are recombinant subunit vaccines, self-assembled as virus like particles from the C-terminal and NANP repeat regions of the circumsporozoite antigen fused to the hepatitis B surface antigen, and then combined with either the adjuvant Matrix-M (i.e. R21/Matrix-M)^1^ or the adjuvant AS01E (i.e. RTS,S/AS01)^2^. These vaccines induce immunity through antibody responses to the circumsporozoite protein (CSP) on the surface of *Plasmodium falciparum* (Pf) sporozoites injected by infected mosquitoes. The passive transfer of anti-CSP antibodies is also partially protective^3^. Sporozoites which evade vaccine-induced humoral immunity invade hepatocytes and give rise to thousands of merozoites, leading to a blood-stage infection. It is unclear why antibody-based protection remains partial despite highly immunogenic vaccines or high concentrations of circulating monoclonal antibodies. Empirically, protection appears to be “leaky” with given antibody levels being inconsistently protective^4^, and with vaccinated individuals experiencing a relative reduction in infection rates but with few individuals completely protected over prolonged follow up at high transmission^5^.

An alternative vaccination strategy is induction of T cells targeting intra-hepatocytic parasites using viral vectors^6^. Previous studies of vaccination with a recombinant chimpanzee adenovirus (ChAd63) then modified Vaccina Ankara (MVA), both encoding the falciparum antigens full length pre-erythrocytic antigen thrombospondin-related adhesive protein (TRAP) fused to a multi-epitope (ME) string (ME-TRAP) have shown efficacy against CHMI among UK adults^7^, no efficacy in West African children^8^, but efficacy in Kenyan adults^9^.

Infectious mosquito bites mostly deliver Pf sporozoites into dermal layers, and sporadically into capillaries ^10^. Direct intravenous injection of sporozoites infects human hosts with 7-fold fewer sporozoites than intradermal injection^11,12^ with many sporozoites failing to reach capillaries^13^. Furthermore, in animal models dermal sporozoites are more readily blocked by anti-CSP antibodies compared with intravenous sporozoites^14,15^.

To our knowledge there are no CHMI studies that compare anti-CSP antibodies against different routes of inoculation of sporozoites in human volunteers. We undertook a CHMI efficacy study in volunteers vaccinated with R21/Matrix-M in Kilifi on the Kenyan Coast, comparing direct intravenous versus intradermal challenge. Furthermore, we re-tested the efficacy of ChAd63/MVA ME-TRAP using only intradermal challenge.

## Results

135 volunteers were screened for eligibility, and 80 were randomized and vaccinated between July 2022 and February 2023. All completed scheduled vaccinations, and the first cohort (i.e. n=40) were eligible for CHMI. Three did not proceed to CHMI due to SARS-CoV-2 positive results (n=2) and increase in liver enzymes (ALT, n=1), so that 37 completed CHMI (Fig. S1). One volunteer was qPCR positive for malaria parasites prior to vaccination, and all were qPCR negative for malaria parasites prior to CHMI, and with a predominance of young male volunteers (Table S1).

### Adverse Events

Following vaccination, self-limiting local solicited adverse events and general adverse events such as headache or fatigue were detected in a minority of volunteers (Table S2). No event was reported as severe and four events were reported as moderate, with a median and maximum duration of 4.5 and 5 days, respectively. During CHMI, there were no immediate or early adverse events, but after day 7 fever, headache and other mild events were common (Table S3). Six events were reported to be moderate and none to be severe. All events resolved on follow up.

Abnormal blood tests, primarily transient elevations in liver enzymes and/or low white blood counts were identified in several volunteers but were not clinically significant, and resolved on follow up (Table S4). There were clinically non-significant minor elevations in liver enzymes among the R21 ID group, and during CHMI there were clinically non-significant, transient abnormalities of liver enzymes and platelet counts (Table S5).

### Immunogenicity

Following the vaccination course with R21, anti-NANP antibodies rose from baseline levels of just under 10 ELISA Units (EU) to above 1000 EU in the R21-vaccinated volunteers but remained below 10 among the control group (fig S2). Peak responses were geometric means of 2,651 and 1,355 and minimum to maximum ranges of 1,200-12,678 vs 590-4,807, for DVI vs ID challenged groups, respectively (p=0.14).

Following vaccination with viral vectors encoding ME-TRAP, spot forming units per million PBMC (sfu) increased from geometric mean of 104 sfu (95%CI 70-153) at baseline to a peak of 735 (95%CI 364-1485) and 335 sfu (95%CI 162-691) on the day of challenge, compared with 174 sfu (95%CI 74-409) among the control group (Figure S3).

Volunteers had evidence of past exposure to malaria as evidenced by pre-vaccination IgG antibodies against schizont extract (Figure S4). Historic controls had higher titres than the newly recruited cohort (geometric means of 1,244,000 EU (95% CI 693,000-2,235,000) vs 394,000 EU (95% CI 258,000-602,000), respectively, *p*=0.0009), and the DVI and ID challenged groups had similar schizont extract antibiotics (geometric means of 284,000 EU (95% CI 158,000-511,000) vs 434,000 EU (95% CI 252,000-745,000) respectively, p=0.4).

### Controlled Human Malaria Infection (CHMI)

CHMI was conducted 28 days after the last vaccination. All eight unvaccinated control volunteers, and all 12 volunteers vaccinated with ME-TRAP, became PCR positive for malaria parasites during CHMI with PfSPZ Challenge by ID, with observed typical parasite growth curves on quantitative PCR. Seven of 8 control volunteers and 11 of 12 ME-TRAP volunteers met criteria for malaria treatment during CHMI, and one control and one ME-TRAP volunteer remained just below the parasitaemia threshold for treatment (Figure 1, Panel A, Table 1). Parasites were genotyped for AMA1, confirming the challenge parasite strain (NF54) in all cases (Table S6.).

**Table 1.**
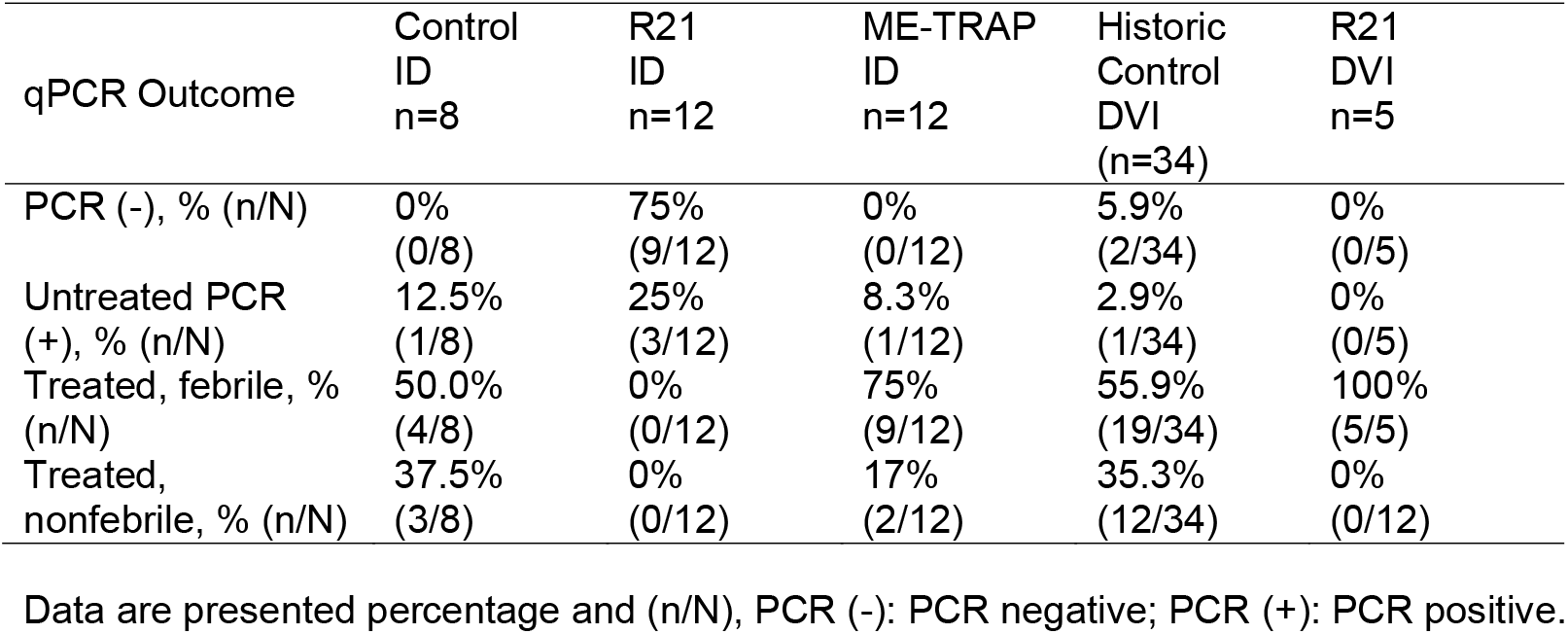
Malaria diagnosis outcome by vaccination and challenge group.

**Figure 1.**
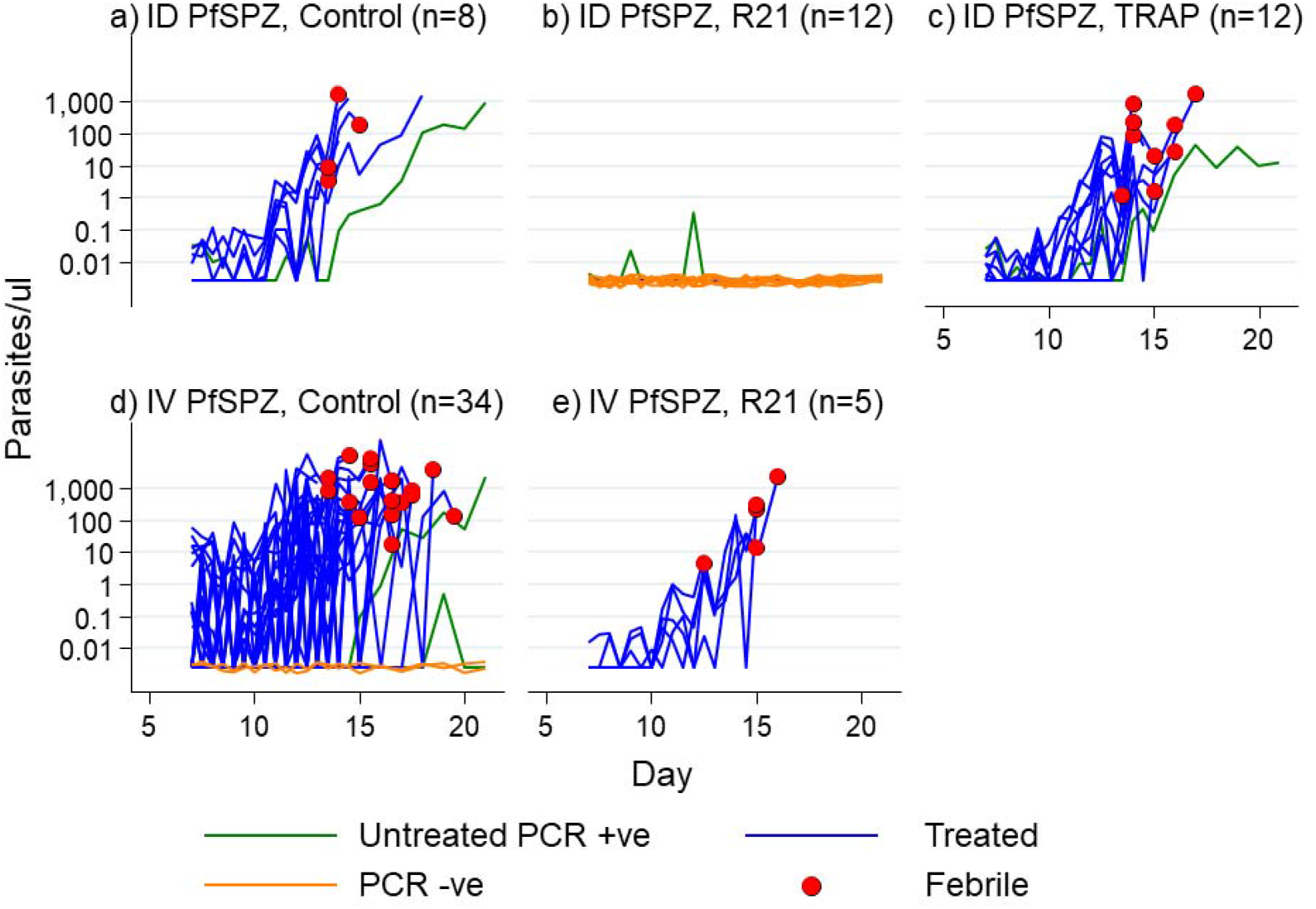
Malaria diagnosis outcome following CHMI. qPCR results (y axis, log transformed), by time after inoculation (x axis) in panels showing (**a**) ID challenge with PfSPZ, control group (n=8); (**b**) ID challenge with PfSPZ, vaccinated with R21/Matrix M; **(c)** ID challenge with PfSPZ, vaccinated with viral vectors encoding ME-TRAP (n=12); (**d**) DVI challenge with PfSPZ, control group (n=34); **(e)** DVI challenge with PfSPZ, vaccinated with R21/Matrix M (n=5). Parasitemia was determined by asexual 18S ribosomal RNA gene qPCR done in Kilifi. Blue lines represent individuals who required reached malaria diagnosis criteria. Green lines represent individuals who did not meet criteria for diagnosis but were qPCR positive. Orange lines represent individuals who were qPCR negative throughout monitoring. Red dots denote individuals who were febrile based on the criteria for diagnosis.

Historical unvaccinated control volunteers who had previously undergone CHMI with PfSPZ Challenge by DVI showed similar growth rates, with 31 of 34 volunteers showing typical growth curves on quantitative PCR and meeting treatment criteria, 2 of 34 being PCR negative and 1 of 34 being PCR positive but not meeting treatment criteria (Figure 1, Panel C).

In contrast, none of the 12 volunteers vaccinated with R21 met criteria for diagnosis following CHMI with PfSPZ Challenge by ID. Three out of the nine volunteers were briefly positive by PCR for malaria parasites but with no evidence of parasite growth hence protected but not sterilely protected (Fig. 1, panel b, Table 1). However, when the 5 volunteers vaccinated with R21 underwent CHMI with PfSPZ Challenge by DVI sporozoites, all 5 showed typical growth curves of parasites, and met diagnosis criteria (Figure 1, Panel D).

The geometric mean parasite densities by PCR were similar over time following PfSPZ Challenge for: the control group by DVI; the control group by ID challenge; for ME-TRAP vaccinees by ID challenge; and for R21 vaccinees by DVI (Figure S5). Hence, parasite inoculum and growth rates appear similar among these four groups.

## Discussion

Our results extend previous observations of protective efficacy by R21 against CHMI delivered by infective mosquito bites in UK adults^16^ and observations of protective efficacy by R21 against natural challenge in the field in West and East African children^1^. Instead of using infectious mosquito bites, we injected PfSPZ Challenge by needle and syringe to compare protective efficacy against direct venous inoculation versus intradermal inoculation of sporozoites.

R21 provided high levels of protective efficacy against PfSPZ Challenge by ID, but we found no evidence of efficacy against PfSPZ Challenge by DVI (i.e. 0 out of 12 volunteers challenged ID met the primary endpoint of requiring treatment vs 5 out of 5 challenged by DVI, p<0.001 by Fisher’s, two-sided). We saw one-off PCR positive signals on a single timepoint among 3 of the 12 volunteers vaccinated with R21 who received PfSPZ Challenge by ID. If we include these volunteers as meeting a secondary endpoint, protection is nevertheless substantial by ID (9 out of 12), and statistically significantly different from DVI (0 out of 5 protected, p=0.009 by Fisher’s, two-sided). Hence, despite the small sample size and the inclusion of 3 low density PCR signals, the variation in protective efficacy of R21 by challenge route was strongly statistically significant.

Was this difference in protective efficacy by challenge route the result of differing PfSPZ doses? PfSPZ Challenge is more infectious when given DVI. To ensure comparable infectivity, we selected PfSPZ Challenge doses that reliably infected unvaccinated volunteers in previous studies (i.e. a 7-fold reduction for DVI at 3,200 PfSPZ Challenge compared with 22,500 cryopreserved PfSPZ Challenge ID ^11,12^. Among the unvaccinated volunteers in our study, neither infection rates (i.e. 8/8 for ID challenge vs 32/34 for IV challenge) nor growth curves (Figure S3) showed variation by challenge route.

We conclude that the different doses resulted in a similar effective inoculum reaching the liver in unvaccinated volunteers, and therefore that the differences seen among R21 vaccinated volunteers imply a qualitatively different interaction between anti-CSP antibodies and PfSPZ Challenge dependent on route of challenge. Furthermore, our data are consistent with animal models suggesting that anti-sporozoite antibodies are more effective against sporozoites in the skin ^14,15^.

Taking together a) that infectious mosquito bites deliver a variable proportion of Pf sporozoites into capillaries or dermal layers ^10^; b) that anti-CSP antibodies correlate noisily with protection against sporozoite infection^4^, with protection appearing leaky^5^; and c) the data presented here that R21 vaccination was ineffective against PfSPZ Challenge by DVI but effective against PfSPZ Challenge by ID injection; we conclude that sporozoites causing infection in the face of high titer anti-CSP antibodies induced by RTS,S or by R21 result from the occasional injection of capillary sporozoites by mosquito bites, producing noise in the correlation between antibody titres and the endpoint, and explaining leaky protection. The infrequent injection of sporozoites into capillaries (i.e. one in five mosquito feeds^10^) is consistent with high levels of efficacy achieved with anti-PfCSP antibody induction despite this lack of efficacy against PfSPZ Challenge by DVI.

More potent or higher titer anti-CSP antibodies may be protective against direct venous injection. In mouse models, high potency monoclonal antibodies are able to inhibit sporozoites in blood whereas lower potency monoclonals can inhibit sporozoites in skin or blood ^17^, and this is further supported by fundings in Saimiri monkeys challenged with vivax sporozoites^18^. Furthermore, vaccines acting through mechanisms apart from anti-CSP antibodies may not exhibit route-dependent protection^19^.

ME-TRAP was not protective against CHMI in our study, and T cell induction judged by ELISpot was lower than that previously seen in European and Kenyan volunteers among whom protective efficacy was observed (i.e. 735 sfu at peak in our study vs 2068 and 1451, respectively)^7,9^. Immunogenicity to this regimen may be suppressed by prior malaria exposure^20^, and may explain variable efficacy.

We relied on historical controls for PfSPZ Challenge by DVI, and prior malaria exposure appears to have been higher in this group (Figure S4). However, the outcomes for DVI vs ID PfSPZ Challenge among R21 vaccinees were based on a contemporaneous and randomized comparison, as was the comparison of R21 vaccinees vs control vaccines for PfSPZ Challenge by ID, and prior malaria exposure did not differ between these groups (fig S4).

Intradermal and direct venous injection of cryopreserved sporozoites may not precisely duplicate infectious mosquito bites, and the latter is likely more representative of natural challenge in the field. For instance, with R21, ID and DVI PfSPZ Challenge would under or overestimate protection in the field, respectively, whereas mosquito bite challenge more accurately predicted subsequent efficacy in the field^16^. On the other hand, our data suggest advantages in studying ID and/or DVI PfSPZ Challenge in making comparisons between products inducing anti-CSP antibodies. The protection of anti-CSP antibodies against these two different routes can be studied in isolation, without the noise in outcome that results from mixed routes with sporadic capillary injection from infectious mosquito bites. For instance, consistent results may be obtained using PfSPZ challenge by ID injection, which can be used to derive antibody thresholds for protection to facilitate immunobridging studies for new products that also induce anti-CSP antibodies. Furthermore, superiority trials between anti-CSP antibody inducing products will be underpowered in classic CHMI studies, but CHMI studies testing higher potency antibodies for protection is against PfSPZ challenge by DVI will be highly powered and sensitive to detect improved antibodies.

## Acknowledgements

We are grateful to all the study volunteers and to the study teams (clinical, nursing, pharmacy, laboratory, and fieldworkers) that have supported the study, including the collaborating teams at Sanaria and the Data Safety Monitoring Board and Local Safety Monitor. We are grateful to the teams at Novavax, Serum Institute India, and Clinical Biomanufacturing Facility at Oxford University for the vaccines. All study team members were notified of the manuscript and reviewed the manuscript. This manuscript is published with permission/approval from KEMRI Director.

## Author Contributions

Conceptualization: MCK, PB, AVSH, SH. Formal analysis: MCK, HK, MM, and PB. Funding acquisition: KJE, AVSH, MH, and PB. Investigation: FO, DK, EK, MSD, DB, JM, ON, HR, SA, MwM, LN, KK, RG, AM, JM, JeM, CN, MoM, JoM, BT, KW, and LOO. Project administration: JW, AL, FRL, and RR. Resources: YA, ERJ, PFB, TLR, BKLS, and SLH. Supervision: MCK, KJE, AVSH, and PB. Writing – original draft: MCK and PB. Writing – review & editing: All coauthors.

## Methods

We undertook a Phase IIb open label, unblinded, randomised single centre study between July 2022 and February 2023 at the KEMRI Wellcome Trust Research Programme in Kilifi Kenya. Prior to commencing activity, approvals were obtained from a National IRB in Kenya and the relevant Oxford IRB (i.e. SERU (KEMRI/SERU/CGMR-C/158/3844) and OxTREC (OxTREC 32-19)) and from the medicines regulatory authority in Kenya (i.e. Pharmacy and Poisons Board (ECCT/19/11/01)). The study was registered with the ClinicalTrials.gov identifier NCT03947190 and with PACTR identifier PACTR202108505632810 to comply with in-country regulatory requirements. Use of Sanaria® PfSPZ Challenge (NF54) is done in accordance with an investigational new drugs application (IND) with the U.S. Food and Drug Administration.

A total of n=56 volunteers were randomized into one of four groups (R21/Matrix M ID, R21/Matrix-M DVI, ChAd63/MVA ME-TRAP ID, and Control ID) across two cohorts to assess the safety, immunogenicity, and protective efficacy in CHMI of R21 adjuvanted with Matrix-M (R21/Matrix-M) in a 0-, 1-, 2-month schedule (Supplementary Methods). The 80 volunteers were divided into 2 cohorts, the first of which was also enrolled into CHMI and the second cohort did not participate in CHMI. For CHMI, we used P. falciparum NF54 sporozoites (Sanaria® PfSPZ Challenge (NF54)) using two alternative inoculation routes: injecting either 22,500 PfSPZ Challenge intradermally (ID) or 3,200 PfSPZ Challenge by direct venous inoculation (DVI).

### Study volunteers and eligibility

Following informed consent, we recruited healthy adult men and women aged between 18-45 years from Kilifi North on the Kenyan Coast, where there is currently very low malaria transmission but with previous low to moderate malaria transmission 16. Clinically significant medical conditions were identified by interview, physical examination, and laboratory screening for renal function, liver function, and serology for HIV, Hepatitis B and C. Volunteers were excluded for clinically significant conditions or blood tests, for likelihood of migration out of the study area, sickle cell disease and trait, pregnancy, and receipt of an investigational product 30 days preceding enrolment.

### Enrollment and Randomization

Volunteers were considered enrolled at the time of randomization, and vaccination was done on the same day. Enrolment took place within 90 days of screening. The volunteers were randomly assigned with randomization via a computer-generated sequence by an independent statistician. A randomization list in the form of password protected spreadsheet was generated using STATA and the data manager set up randomization in REDCap. Study clinicians only clicked to randomize after confirmation of eligibility criteria and immediately prior to vaccination for each volunteer, after which REDCap would reveal the volunteers’ randomization arm that could not be edited. The volunteers were randomized to either one of four groups.

### Vaccines and vaccination

R21 is a pre-erythrocytic protein-in-adjuvant malaria vaccine candidate. It is adjuvanted with Matrix-M™ based on the circumsporozoite protein (CSP) produced by using recombinant Hepatitis B surface antigen (HBsAg) particles expressing the central NANP repeat and the C-terminus1–4. As previously described, volunteers received three vaccinations with R21 at four-week intervals. R21 was thawed to room temperature then mixed with Matrix-M™ before administration (10 μg mixed with Matrix-M 50 μg) and administered intramuscularly.

The viral vectors used were ChAd63 ME-TRAP and MVA ME-TRAP. Volunteers received a single intramuscular dose of each vaccine which was given, sequentially, 8 weeks apart. Both vaccines were thawed to room temperature before administration.

### Controlled human malaria infection (CHMI)

CHMI was undertaken four weeks after the final vaccination. Volunteers were tested for malaria blood stage infection by qPCR and tested for SARS-CoV-2 infection by RT-PCR on naso-pharyngeal swab. Volunteers positive for either were excluded from proceeding to CHMI.

The CHMI agent (i.e. Sanaria® PfSPZ Challenge (NF54)) in cryovials was thawed by partial submersion of each vial for 30 s in a 37±1°C water bath. Trained study staff prepared, diluted, and dispensed PfSPZ Challenge to clinical staff using the diluents phosphate buffered saline (PBS) and 25% human serum albumin (HSA) as previously described within 30 minutes of thawing <5,6>. Challenge was administered in a volume of 0.5 mL using a needle and syringe by direct venous inoculation (DVI) at the standard dose of 3.200 PfSPZ Challenge. Intradermal (ID) injections were administered in two separate syringes in both left and right arms with each syringe containing 11,250 PfSPZ Challenge in 0.05 mL (i.e. 22,500 in total). The injection sites were covered with a sterile dressing, removed no earlier than 1 hour after inoculation.

Study endpoint criteria following CHMI for treatment and/or malaria diagnosis was as previously described ^21–23^, i.e. reaching parasitaemia threshold of 500 parasites/μL and/or presence of important clinical symptoms. Volunteers who reached day 22 were treated without meeting the endpoint.

### Safety assessments

Solicited adverse events were recorded for 7 days after each vaccination, volunteers were assessed within 1 hour of vaccination on the days of vaccination. Volunteers were provided with diary cards, rulers and thermometers and trained on measurement and recording of cutaneous reactions or swelling, and auxiliary temperature. Clinicians telephoned vaccinees daily for 7 days to remind them to record solicited events, and to record and assess unsolicited adverse events. Further in person examination was organized if necessary. Any unsolicited adverse events occurring between 7 and 30 days of each vaccination were recorded based on recall at 4 weeks. Serious adverse events were collected throughout the study. Causality was assessed by clinicians.

### Immunology

Plasma samples were separated from whole blood for serology. We conducted ELISAs for levels of anti-NANP antibody and anti-whole schizont antibody (3D7 strain) as previously described ^24^ PBMC were separated by density gradient centrifugation from heparinized whole blood. Ex vivo ELISpot assays were performed over 18 hours of incubation using Multiscreen IP ELISPOT plates (Millipore, Watford, UK) and Mabtech IFNγ SA-ALP antibody kits (Mabtech, Nacka, Sweden). Peptide pools were tested in duplicate with 250,000 PBMC per well. TRAP peptides were 20 amino acids in length, overlapping by 10 amino acids (ProImmune, UK).

Responses were averaged across duplicates, background from negative control wells was subtracted, and summed across TRAP pools plus ME. Plates were counted using an AID automated ELISPOT counter (AID Diagnostika GmbH, Strassberg, Germany algorithm C), using identical settings for all plates. The lower limit of detection for the assay was 20 SFU. *qPCR for parasite detection*

Venous blood was collected twice daily from days 8 to 15 after PfSPZ Challenge inoculation and then daily from days 16 to 22 for the detection of the 18 S ribosomal RNA *P. falciparum* gene using qPCR: in triplicates in a TaqMan assay using primers and probes previously described^23,25^. Non-template water (non-template control, NTC) was used as a negative control and cultured parasites of known quantity used as positive control, with sample parasite quantification undertaken against DNA extracted from known cultured parasite standards using 8 serial dilutions. Standard curves were checked against the WHO external quantified quality control sample^23,25^.

### Parasite genotyping

Parasite genotyping targeted the ama1 (PF3D7_1133400) gene using amplicon deep sequencing as previously described9. Briefly, ama1 amplicons spanning nucleotides 441– 946 were generated from each sample, in duplicate, using primers designed in a previous study <7>. Deep sequencing of the sequencing library was performed on the SpotON FlowCell R10.4.1 (Oxford Nanopore Technologies, catalogue number FLOW-MIN114-1) at the KEMRI-Wellcome Trust Laboratories. Sequence data analysis was performed in SeekDeep version 3.0.110, as previously described ^26^. ama1 microhaplotypes with fewer than 250 reads or those detected at less than 5% minor allele frequency were discarded unless the microhaplotype was independently detected in additional samples at >5% frequency.

### Statistical analysis

The intent-to-treat (ITT) cohort included volunteers receiving one or more vaccines. The according-to-protocol (ATP) cohort included all evaluable volunteers meeting eligibility criteria and complied with study procedures. Safety is reported ITT, immunogenicity is reported ATP and efficacy in CHMI is reported ITT for the cohort undergoing CHMI. Safety data were tabulated descriptively, immunogenicity was described by geometric means with 95% confidence intervals. CHMI data were reported by description of quantitative PCR over time, then by frequency of outcome categories (as previously defined in Kapulu et al 2016) according to vaccine group.

## Data Availability Statement

Data are available through the online repository for KEMRI-Wellcome Trust Research Programme: Harvard Dataverse: https://doi.org/10.7910/DVN/TNHS14

## Code Availability Statement

Not applicable

## References (Main Text)

1. Datoo, M. S. et al. Safety and efficacy of malaria vaccine candidate R21/Matrix-M in African children: a multicentre, double-blind, randomised, phase 3 trial. The Lancet 403, 533–544 (2024).

2. Clinical Trials Partnership, S. Efficacy and safety of RTS,S/AS01 malaria vaccine with or without a booster dose in infants and children in Africa: Final results of a phase 3, individually randomised, controlled trial. The Lancet 386, 31–45 (2015).

3. Kayentao, K. et al. Subcutaneous Administration of a Monoclonal Antibody to Prevent Malaria. New England Journal of Medicine 390, 1549–1559 (2024).

4. Olotu, A. I., Fegan, G. & Bejon, P. Further analysis of correlates of protection from a phase 2a trial of the falciparum malaria vaccines RTS,S/AS01B and RTS,S/AS02A in malaria-naive adults. Journal of Infectious Diseases 201, 970–971 (2010).

5. Olotu, A. et al. Four-Year Efficacy of RTS,S/AS01E and Its Interaction with Malaria Exposure. New England Journal of Medicine 368, 1111–1120 (2013).

6. Kimani, D. et al. Translating the Immunogenicity of Prime-boost Immunization With ChAd63 and MVA ME-TRAP From Malaria Naive to Malaria-endemic Populations. Molecular Therapy, Vol 22, Issue 11, pp. 1992-2003. 22, 1992–2003 (2014).

7. Hodgson, S. H. et al. Evaluation of the Efficacy of ChAd63-MVA Vectored Vaccines Expressing Circumsporozoite Protein and ME-TRAP Against Controlled Human Malaria Infection in Malaria-Naive Individuals. J Infect Dis 211, 1076–1086 (2015).

8. Tiono, A. B. et al. First field efficacy trial of the ChAd63 MVA ME-TRAP vectored malaria vaccine candidate in 5-17 months old infants and children. PLoS One 13, e0208328 (2018).

9. Ogwang, C. et al. Prime-boost vaccination with chimpanzee adenovirus and modified vaccinia Ankara encoding TRAP provides partial protection against Plasmodium falciparum infection in Kenyan adults. Sci Transl Med 7, (2015).

10. Ponnudurai, T., Lensen, A. H. W., Gemert, G. J. A., Bolmer, M. G. & Meuwissen, J. H. T. Feeding behaviour and sporozoite ejection by infected Anopheles stephensi. Trans R Soc Trop Med Hyg 85, 175–180 (1991).

11. Shekalaghe, S. et al. Controlled human malaria infection of Tanzanians by intradermal injection of aseptic, purified, cryopreserved plasmodium falciparum sporozoites. American Journal of Tropical Medicine and Hygiene 91, 471–480 (2014).

12. Mordmüller, B. et al. Direct venous inoculation of Plasmodium falciparum sporozoites for controlled human malaria infection: a dose-finding trial in two centres. Malar J 14, 117 (2015).

13. Hopp, C. S. et al. Longitudinal analysis of Plasmodium sporozoite motility in the dermis reveals component of blood vessel recognition. Elife 4, (2015).

14. Flores-Garcia, Y. et al. Antibody-Mediated Protection against Plasmodium Sporozoites Begins at the Dermal Inoculation Site. mBio 9, (2018).

15. Aliprandini, E. et al. Cytotoxic anti-circumsporozoite antibodies target malaria sporozoites in the host skin. Nat Microbiol 3, 1224–1233 (2018).

16. Venkatraman, N. et al. R21 in Matrix-M adjuvant in UK malaria-naive adult men and non-pregnant women aged 18–45 years: an open-label, partially blinded, phase 1–2a controlled human malaria infection study. Lancet Microbe 0, 100867 (2024).

17. Aguirre-Botero, M. C. et al. Cytotoxicity of human antibodies targeting the circumsporozoite protein is amplified by 3D substrate and correlates with protection. Cell Rep 42, (2023).

18. Charoenvit, Y. et al. Inability of malaria vaccine to induce antibodies to a protective epitope within its sequence. Science 251, 668–671 (1991).

19. Roozen, G. V. T. et al. Single immunization with genetically attenuated PfΔmei2 (GA2) parasites by mosquito bite in controlled human malaria infection: a placebo-controlled randomized trial. Nature Medicine 2025 1–5 (2025) doi:10.1038/s41591-024-03347-2.

20. Bejon, P. et al. The Induction and Persistence of T Cell IFN-γ Responses after Vaccination or Natural Exposure Is Suppressed by Plasmodium falciparum. The Journal of Immunology 179, 4193–4201 (2007).

21. Kapulu, M. C. et al. Safety and PCR monitoring in 161 semi-immune Kenyan adults following controlled human malaria infection. JCI Insight 6, (2021).

22. Kapulu, M. C. et al. Controlled human malaria infection (CHMI) outcomes in Kenyan adults is associated with prior history of malaria exposure and anti-schizont antibody response. BMC Infect Dis 22, (2022).

23. Kibwana, E. et al. Quantification of Plasmodium falciparum: validation of quantitative polymerase chain reaction assays for detection of parasites in controlled human malaria infection studies. Frontiers in Malaria 3, 1497613 (2025).

24. Sang, S. et al. Safety and immunogenicity of varied doses of R21/Matrix-MTM vaccine at three years follow-up: A phase 1b age de-escalation, dose-escalation trial in adults, children, and infants in Kilifi-Kenya. Wellcome Open Res 8, (2023).

25. Kapulu, M. C. et al. Safety and PCR monitoring in 161 semi-immune Kenyan adults following controlled human malaria infection. JCI Insight 6, (2021).

26. Wamae, K. et al. Targeted amplicon deep sequencing of ama1 and mdr1 to track within-host P. falciparum diversity throughout treatment in a clinical drug trial. Wellcome Open Res 7, 95 (2024).

## References (Methods)

1. Kapulu, M. C. et al. Safety and PCR monitoring in 161 semi-immune Kenyan adults following controlled human malaria infection. JCI Insight 6, (2021).

2. Kapulu, M. C. et al. Controlled human malaria infection (CHMI) outcomes in Kenyan adults is associated with prior history of malaria exposure and anti-schizont antibody response. BMC Infect Dis 22, (2022).

3. Sang, S. et al. Safety and immunogenicity of varied doses of R21/Matrix-MTM vaccine at three years follow-up: A phase 1b age de-escalation, dose-escalation trial in adults, children, and infants in Kilifi-Kenya. Wellcome Open Res 8, (2023).

4. Wamae, K. et al. Targeted amplicon deep sequencing of ama1 and mdr1 to track within-host P. falciparum diversity throughout treatment in a clinical drug trial. Wellcome Open Res 7, 95 (2024).

