## Supplementary Appendix for "R21/Matrix-M Protects Against Dermal but not Against Venous Parasites in Human Challenge Studies"

### Supplementary Tables and Figures for R21/Matrix-M Protects against Dermal but not against Venous Parasites in Human Challenge Studies

#### Table S1: Baseline Characteristics Based on Vaccine Enrolment and CHMI Group.

| Challenged | | | | |  | Not Challenged | | | Total  (N=80) | Historic Control DVI  (n=34) |
| --- | --- | --- | --- | --- | --- | --- | --- | --- | --- | --- |
|  | R21 ID  (n=12) | R21 DVI  (n=7) | ME-TRAP  ID  (n=12) | Control ID  (n=9) | R21 ID  (n=12) | R21 DVI  (n=7) | ME-TRAP  ID  (n=12) | Control ID  (n=9) |  |  |
| Age in years, mean (SD) | 28.17 (6.1) | 26.14 (4.1) | 28.83  (5.1) | 30.78 (8.1) | 27.42 (6.8) | 25.43 (6.0) | 28.08 (7.1) | 26.67 (3.5) | 28 (6) | 28 (8) |
| Male, % (n/N) | 83.3% (10/12) | 71.4% (5/7) | 66.7% (8/12) | 55.6% (5/9) | 83.3% (10/12) | 71.4% (5/7) | 58.3% (7/12) | 77.8% (7/9) | 71% (57/80) | 79% (27/34) |
| Female, % (n/N) | 16.7% (2/12) | 28.6% (2/7) | 33.3% (4/12) | 44.4% (4/9) | 16.7% (2/12) | 28.6% (2/7) | 41.7% (5/12) | 22.2% (2/9) | 29% (23/80) | 21% (7/34) |
| BMI, kg/m2, mean (SD) | 21.69 (1.7) | 19.72 (1.8) | 23.88  (4.8) | 20.55 (1.9) | 21.74 (2.4) | 20.71 (1.8) | 23.29 (3.9) | 19.79 (1.5) | 21.7 (3.1) | 31.0 (3.5) |

Data are presented either as mean (SD), percentage and (n/N), or median and (min-max). n=number of healthy volunteers enrolled to each group. BMI, Body mass index.

#### Table S2. Local and general adverse events post-vaccination.

|  | **Day 0** | | | |
| --- | --- | --- | --- | --- |
| **Local AEs** | R21 ID  n=24 | R21 DVI  n=14 | ME-TRAP ID  N=24 | Total  N=62 |
| Pain at injection site | 1 (4.2%) | 0 | 2(8.3%) | 3(4.8%) |
| Induration | 0 | 0 | 0 | 0 |
| Erythema | 0 | 0 | 0 | 0 |
|  | **General AEs** | | | |
| Arthralgia | 0 | 0 | 1(4.2%) | 1(4.2) |
| Chills | 0 | 0 | 1(4.2%) | 1(4.2) |
| Fatigue | 2 (8.3%) | 0 | 3(12%) | 5(8.1%) |
| Fever | 0 | 1 (7.1%) | 1(4.2%) | 2(3.2) |
| Headache | 0 | 1 (7.1%) | 5(21%) | 6(9.7%) |
| Malaise | 2 (8.3%) | 0 | 2(8.3%) | 4(6.5%) |
| Myalgia | 0 | 0 | 1(4.2%) | 1(4.2%) |
|  | **Day 28** | | | |
| **Local AEs** | R21 ID  n=24 | R21 DVI  n=14 |  | Total  N=38 |
| Pain at injection site | 0 | 0 |  | 0 |
| Induration | 0 | 0 |  | 0 |
| Erythema | 0 | 0 |  | 0 |
|  | General AEs | | | |
| Arthralgia | 0 | 0 |  | 0 |
| Chills | 1 (4.2%) | 1 (7.1%) |  | 2 (5.3%) |
| Fatigue | 0 | 1 (7.1%) |  | 1 (2.6%) |
| Fever | 1 (4.2%) | 1 (7.1%) |  | 2 (5.3%) |
| Headache | 0 | 1 (7.1%) |  | 1 (2.6%) |
| Malaise | 0 | 0 |  | 0 |
| Myalgia | 1 (4.2%) | 2 (14%) |  | 3 (7.9%) |
|  | **Day 56** | | | |
| **Local AEs** | R21 ID  n=24 | R21 DVI  n=14 | ME-TRAP ID  N=24 | Total  N=62 |
| Pain at injection site | 0 | 0 | 4(16.7%) | 4(6.5%) |
| Induration | 0 | 0 | 1(4.2%) | 1(1.6%) |
| Erythema | 0 | 0 | 0 | 0 |
|  | **General AEs** | | | |
| Arthralgia | 0 | 0 | 1(4.2%) | 1(1.6%) |
| Chills | 0 | 0 | 1(4.2%) | 1(1.6%) |
| Fatigue | 0 | 0 | 1(4.2%) | 1(1.6%) |
| Fever | 1 (4.2%) | 0 | 5(20.8%) | 6(9.7) |
| Headache | 0 | 2 (14%) | 8(33.3%) | 10(16.1%) |
| Malaise | 0 | 0 | 3(12.5%) | 3(4.8%) |
| Myalgia | 1(4.2) | 0 | 3(12.5%) | 4(6.5%) |

Data are presented as n (%). Percentages indicate volunteers with adverse events over the total number of volunteers per vaccination group. All adverse events are reported based on maximum severity over the follow-up period. The median duration of events is 1 day.

#### Table S3. General adverse events between C+8-DoT and DoT-Exit.

|  | | **C+8-DoT*** | | | | |
| --- | --- | --- | --- | --- | --- | --- |
| **Systemic solicited** | **R21**  **ID**  **n=12** | | **R21**  **DVI**  **n=5** | **ME-TRAP**  **ID**  **n=12** | **Control**  **ID**  **n=8** | **Total**  **N=37** |
| Fever | 1 (8.3) | | 5 (100) | 11 (91.7) | 5 (62.5) | 22 (59.5) |
| Headache | 2 (16.7) | | 4 (80) | 8 (66.7) | 3 (37.5) | 17 (45.9) |
| Fatigue | 1 (8.3) | | 2 (40) | 5 (41.7) | 4 (50) | 12 (32.0) |
| Low back pain | 0 | | 2 (40) | 5 (41.7) | 3 (37.5) | 10 (27.0) |
| Myalgia | 1 (8.3) | | 1 (20) | 3 (25) | 2 (25) | 7 (18.9) |
| Arthralgia | 0 | | 1 (20) | 2 (16.7) | 3 (37.5) | 6 (16.2) |
| Chills | 0 | | 2 (40) | 2 (16.7) | 2 (25) | 6 (16.2) |
| Anorexia | 0 | | 2 (40) | 2 (16/7) | 2 (25) | 5 (13.5) |
| Nausea | 1 (8.3) | | 0 | 0 | 2 (25) | 3 (8.1) |
| Sweating | 0 | | 0 | 1 (8.3) | 2 (25) | 3 (8.1) |
| Rigors | 0 | | 0 | 0 | 0 | 0 |
| Vomiting | 0 | | 0 | 0 | 0 | 0 |
|  | | **Systemic unsolicited** | | | | |
| Neck pain | 0 | | 0 | 1 (8.3) | 1 (12.5) | 2 (5.4) |
| Abdominal pain | 1 (8.3) | | 1 (20) | 1 (8.3) | 0 | 3 (8.1) |
|  | | **DoT – Exit**** | | | | |
| **Systemic solicited** | **R21**  **ID**  **n=12** | | **R21**  **DVI**  **n=5** | **ME-TRAP**  **ID**  **N=12** | **Control**  **ID**  **n=8** | **Total**  **N=37** |
| Fever | 0 | | 3 (60) | 2 (16.7) | 2 (25) | 7 (18.9) |
| Headache | 1 (8.3) | | 3 (60) | 3 (25) | 4 (50) | 11 (29.7) |
| Fatigue | 2 (16.7) | | 1 (20) | 2 (16.7) | 4 (50) | 9 (24.3) |
| Myalgia | 0 | | 1 (20) | 4 (33.3) | 3 (37.5) | 8 (21.6) |
| Low back pain | 0 | | 0 | 2 (16.7) | 2 (25) | 4 (10.8) |
| Arthralgia | 0 | | 1 (20) | 2 (16.7) | 3 (37.5) | 6 (16.2) |
| Anorexia | 0 | | 1 (20) | 1 (8.3) | 2 (25) | 4 (10.8) |
| Nausea | 0 | | 0 | 1 (8.3) | 3 (37.5) | 4 (10.8) |
| Chills | 0 | | 1 (20) | 1 (8.3) | 1 (12.5) | 3 (8.1) |
| Sweating | 0 | | 0 | 0 | 2 (25) | 2 (5.4) |
| Rigors | 0 | | 0 | 0 | 0 | 0 |
| Vomiting | 0 | | 0 | 0 | 0 | 0 |
|  | | **System unsolicited** | | | | |
| Cough | 0 | | 0 | 1 (8.3) | 0 | 1 (2.7) |
| Mouth sores | 0 | | 0 | 2 (16.7) | 0 | 2 (5.4) |
| Tonsilitis | 0 | | 0 | 0 | 1 (12.5) | 1 (2.7) |

Data are presented as n (%). Percentages indicate volunteers with adverse events over the total number of volunteers per vaccination group. All adverse events are reported based on maximum severity over the follow-up period. The median duration of events is 1 day. *C+8 to DoT refers to the timeframe starting from 7 days after a challenge to the day of diagnosis. **DoT to Exit refers to period of treatment from the diagnosis to inpatient exit.

#### Table S4. Volunteers with out-of-range safety blood results during vaccination (n=62).

| **Parameter** | **Gender** | **Limit** | **Treatment Group** | **Timepoint** | | |
| --- | --- | --- | --- | --- | --- | --- |
|  | | | | **D28** | **D56** | **C-1** |
| **Chemistry** | | | |  |  |  |
| Creatinine | Male | >115 | R21 ID | 0 | 0 | 0 |
|  |  |  | R21 DVI | 1 | 1 | 0 |
|  |  |  | ME-TRAP ID | 0 | 1 | 0 |
|  |  |  | Control ID | 0 | 0 | 1 |
|  | Female | >91 | R21 ID | 0 | 0 | 0 |
|  |  |  | R21 DVI | 1 | 0 | 0 |
|  |  |  | ME-TRAP ID | 0 | 1 | 1 |
|  |  |  | Control ID | 0 | 0 | 0 |
| AST | Male | >47 | R21 ID | 4 | 3 | 5 |
|  |  |  | R21 DVI | 0 | 2 | 1 |
|  |  |  | ME-TRAP ID | 0 | 0 | 4 |
|  |  |  | Control ID | 0 | 0 | 1 |
|  | Female | >42 | R21 ID | 0 | 0 | 1 |
|  |  |  | R21 DVI | 0 | 0 | 1 |
|  |  |  | ME-TRAP ID | 0 | 1 | 3 |
|  |  |  | Control ID | 0 | 0 | 1 |
| ALT | Male | >35 | R21 ID | 6 | 4 | 10 |
|  |  |  | R21 DVI | 1 | 3 | 2 |
|  |  |  | ME-TRAP ID | 0 | 2 | 8 |
|  |  |  | Control ID | 0 | 0 | 5 |
|  | Female | >35 | R21 ID | 1 | 0 | 3 |
|  |  |  | R21 DVI | 0 | 0 | 1 |
|  |  |  | ME-TRAP ID | 0 | 1 | 3 |
|  |  |  | Control ID | 0 | 0 | 1 |
| **Haematology** | | | | | | |
| Hb | Male | <11.5 | R21 ID | 0 | 0 | 0 |
|  |  |  | R21 DVI | 0 | 0 | 0 |
|  |  |  | ME-TRAP ID | 0 | 0 | 0 |
|  |  |  | Control ID | 0 | 0 | 0 |
|  | Female | <9.5 | R21 ID | 0 | 0 | 0 |
|  |  |  | R21 DVI | 0 | 0 | 0 |
|  |  |  | ME-TRAP ID | 0 | 0 | 1 |
|  |  |  | Control ID | 0 | 0 | 0 |
| WBC (Total) | Male | <3.3 0R >8.4 | R21 ID | 6 | 6 | 1 |
|  |  |  | R21 DVI | 3 | 3 | 0 |
|  |  |  | ME-TRAP ID | 0 | 1 | 1 |
|  |  |  | Control ID | 0 | 0 | 1 |
|  | Female | <3.3 OR >8.2 | R21 ID | 0 | 0 | 0 |
|  |  |  | R21 DVI | 0 | 0 | 0 |
|  |  |  | ME-TRAP ID | 0 | 0 | 2 |
|  |  |  | Control ID | 0 | 0 | 0 |
| Platelets | Male | <124 OR >358 | R21 ID | 0 | 0 | 1 |
|  |  |  | R21 DVI | 1 | 1 | 0 |
|  |  |  | ME-TRAP ID | 0 | 0 | 1 |
|  |  |  | Control ID | 0 | 0 | 0 |
|  | Female | <127 OR >397 | R21 ID | 0 | 1 | 1 |
|  |  |  | R21 DVI | 0 | 0 | 0 |
|  |  |  | ME-TRAP ID | 0 | 0 | 0 |
|  |  |  | Control ID | 0 | 0 | 1 |

ALT denotes alanine transaminase; AST aspartate aminotransferase; D Day after vaccination; Hb, Haemoglobin; WBC white blood count.

#### Table S5. Volunteers with out-of-range safety blood results during CHMI (n=37).

| **Parameter** | **Gender** | **Limit** | **Timepoint** | | | | | |
| --- | --- | --- | --- | --- | --- | --- | --- | --- |
|  |  |  | C+6 | C+8 | C+10 | DoT | C+36 | C+91 |
| **Chemistry** | | | | | | | | |
| Creatinine | Male | >115 | 1 | 0 | 0 | 4 | 0 | 0 |
|  | Female | >91 | 0 | 0 | 0 | 2 | 0 | 0 |
| AST | Male | >47 | 2 | 2 | 3 | 7 | 0 | 0 |
|  | Female | >42 | 2 | 2 | 2 | 2 | 0 | 0 |
| ALT | Male | >35 | 11 | 11 | 11 | 14 | 0 | 0 |
|  | Female | >35 | 4 | 4 | 4 | 3 | 0 | 0 |
| **Haematology** | | | | | | | | |
| Hb | Male | <11.5 | 0 | 0 | 0 | 0 | 0 | 0 |
|  | Female | <9.5 | 0 | 0 | 0 | 0 | 0 | 0 |
| WBC (Total) | Male | <3.3 OR >8.4 | 0 | 0 | 0 | 2 | 3 | 2 |
|  | Female | <3.2 OR >8.2 | 0 | 2 | 0 | 0 | 0 | 2 |
| Platelets | Male | <124 OR >358 | 0 | 1 | 0 | 4 | 3 | 0 |
|  | Female | <127 OR >397 | 0 | 1 | 0 | 0 | 2 | 0 |

ALT denotes alanine transaminase; AST aspartate aminotransferase; C+ day after challenge; Hb, Haemoglobin; WBC white blood count.

#### Table S6. AMA1 parasite sequence genotype reads for volunteers with detectable parasites.

| Sample ID | Timepoint | Target gene | Genotype target | Read count 1 | Read count 2 |
| --- | --- | --- | --- | --- | --- |
| 22KV054 | DoT | PFAMA1 | PF3D7/PFNF54 | 19,708 | 27,490 |
| 22KV055 | DoT | PFAMA1 | PF3D7/PFNF54 | 22,686 | 21,885 |
| 22KV058 | DoT | PFAMA1 | PF3D7/PFNF54 | 16,989 | 26,223 |
| 22KV060 | DoT | PFAMA1 | PF3D7/PFNF54 | 18,654 | 21,866 |
| 22KV063 | DoT | PFAMA1 | PF3D7/PFNF54 | 17,001 | 28,095 |
| 22KV064 | C+22 | PFAMA1 | PF3D7/PFNF54 | 13,677 | 3,857 |
| 22KV068 | DoT | PFAMA1 | PF3D7/PFNF54 | 22,695 | 37,502 |
| 22KV070 | DoT | PFAMA1 | PF3D7/PFNF54 | 377 | 25,368 |
| 22KV073 | DoT | PFAMA1 | PF3D7/PFNF54 | 17,196 | 14,649 |
| 22KV074 | DoT | PFAMA1 | PF3D7/PFNF54 | 55,486 | 76,725 |
| 22KV075 | DoT | PFAMA1 | PF3D7/PFNF54 | 19,772 | 19,288 |
| 22KV077 | DoT | PFAMA1 | PF3D7/PFNF54 | 26,164 | 17,146 |
| 22KV081 | DoT | PFAMA1 | PF3D7/PFNF54 | 14,934 | 20,812 |
| 22KV083 | DoT | PFAMA1 | PF3D7/PFNF54 | 20,060 | 94,730 |
| 22KV084 | DoT | PFAMA1 | PF3D7/PFNF54 | 12,950 | 33,903 |
| 22KV085 | DoT | PFAMA1 | PF3D7/PFNF54 | 14,295 | 21,380 |
| 22KV090 | DoT | PFAMA1 | PF3D7/PFNF54 | 20,623 | 19,108 |
| 22KV091 | DoT | PFAMA1 | PF3D7/PFNF54 | 18,953 | 15,031 |
| 22KV094 | DoT | PFAMA1 | PF3D7/PFNF54 | 26,501 | 17,762 |
| 22KV096 | DoT | PFAMA1 | PF3D7/PFNF54 | 19,543 | 21,581 |
| 22KV100 | C+22 | PFAMA1 | PF3D7/PFNF54 | 23,580 | 27,981 |
| 22KV103 | DoT | PFAMA1 | PF3D7/PFNF54 | 15,594 | 10,257 |
| 22KV108 | DoT | PFAMA1 | PF3D7/PFNF54 | 16,517 | 18,487 |
| 3D7 Control | N/A | PFAMA1 | PF3D7/PFNF54 | 23,384 | 30,708 |

C+ denotes day after challenge; DoT, day of treatment; N/A not applicable. Read counts 1 and 2 represent the *ama1* sequence read depths obtained from the two duplicate PCR aliquots of the original sample.

#### Figure S1. Study Flow Diagram.

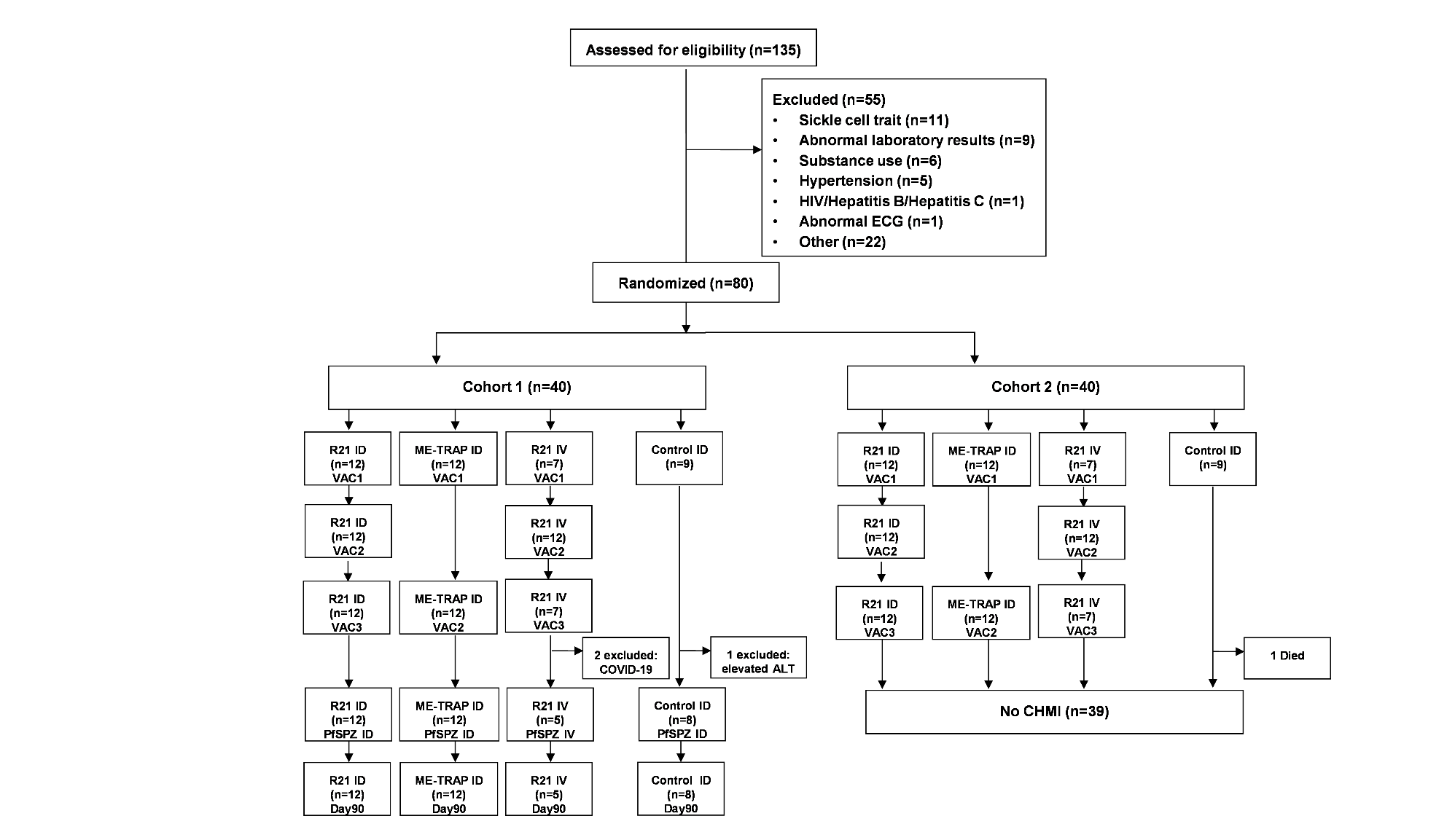

Out of one-hundred and thirty-five volunteers screened, fifty-six volunteers who were eligible and met the enrolment criteria were randomized into one of four groups across two enrolment cohorts 4 weeks apart. Of the 55 excluded volunteers those with abnormal laboratory results, included low hemoglobin levels (<10g/dl for females (n=6)); thrombocytopenia (n=2); and elevated levels of ALTs (n=1). For other exclusions, these included being eligible but: did not turn up for screening results (n=4); not on any method of contraception (n=3); number required attained (n=3); did not meet location of residence criteria (n=2); and did not attend enrolment visit (n=2). Other reasons with frequency of 1 (n=8) were: written informed consent not signed; unstable blood pressure values; positive pregnancy test; abdominal hernia, neurofibromatosis, allergy to ibuprofen, failed Test of Understanding (TOU), history of schizophrenia. 7 volunteers had more than one screening failure and these were elevated laboratory values (n=4), abdominal mass (n=1), abnormal ECG (n=1), and substance use (n=1).

#### Figure S2. NANP Vaccine-induced Antibody Responses over Time.

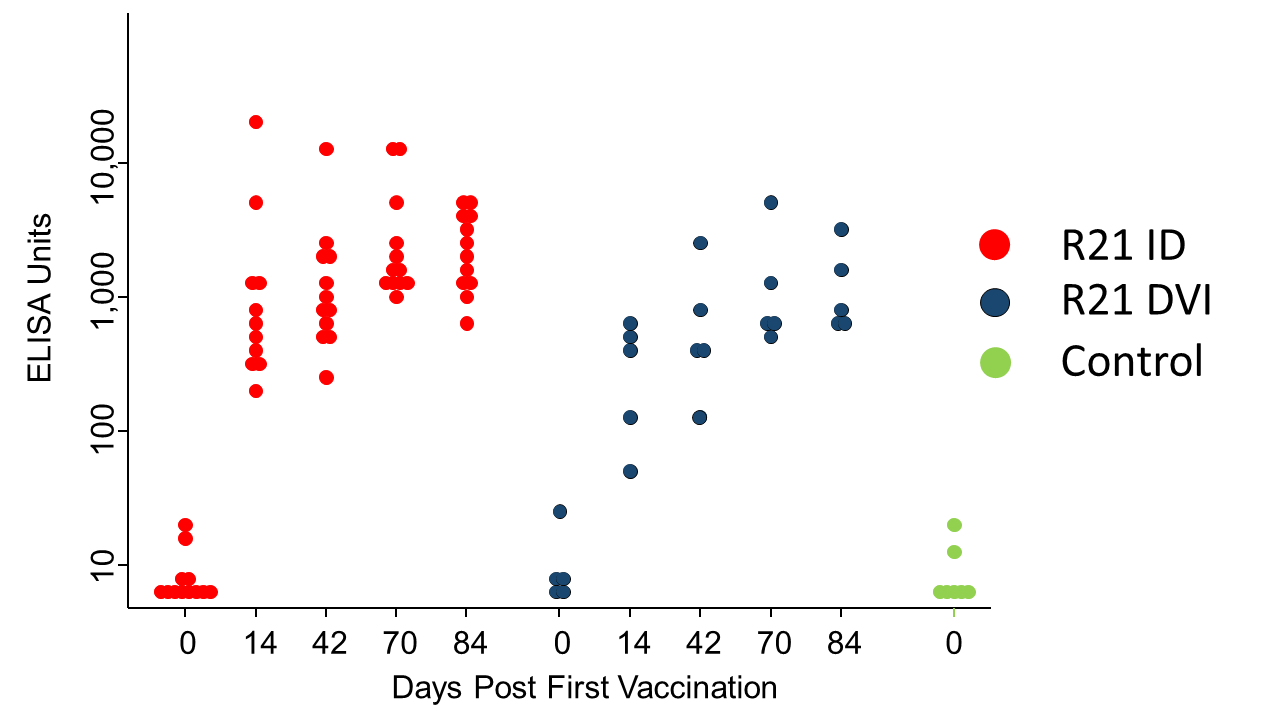

NANP IgG ELISA units are shown over time with individual scatter dot plots per vaccination group.

#### Figure S3. ME-TRAP Vaccine-induced T Cell Responses over Time.

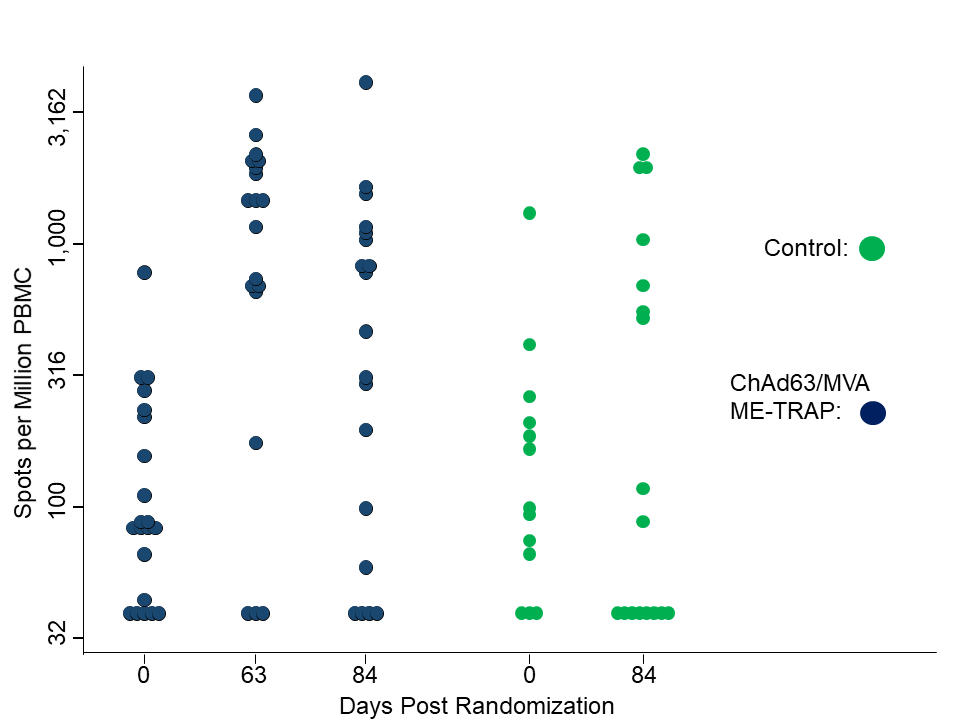

ME-TRAP IFNγ positive spots by ELISpot per million PBMC are shown over time with individual scatter dot plots per group and time point.

#### Figure S4. Past Exposure Antibody Data

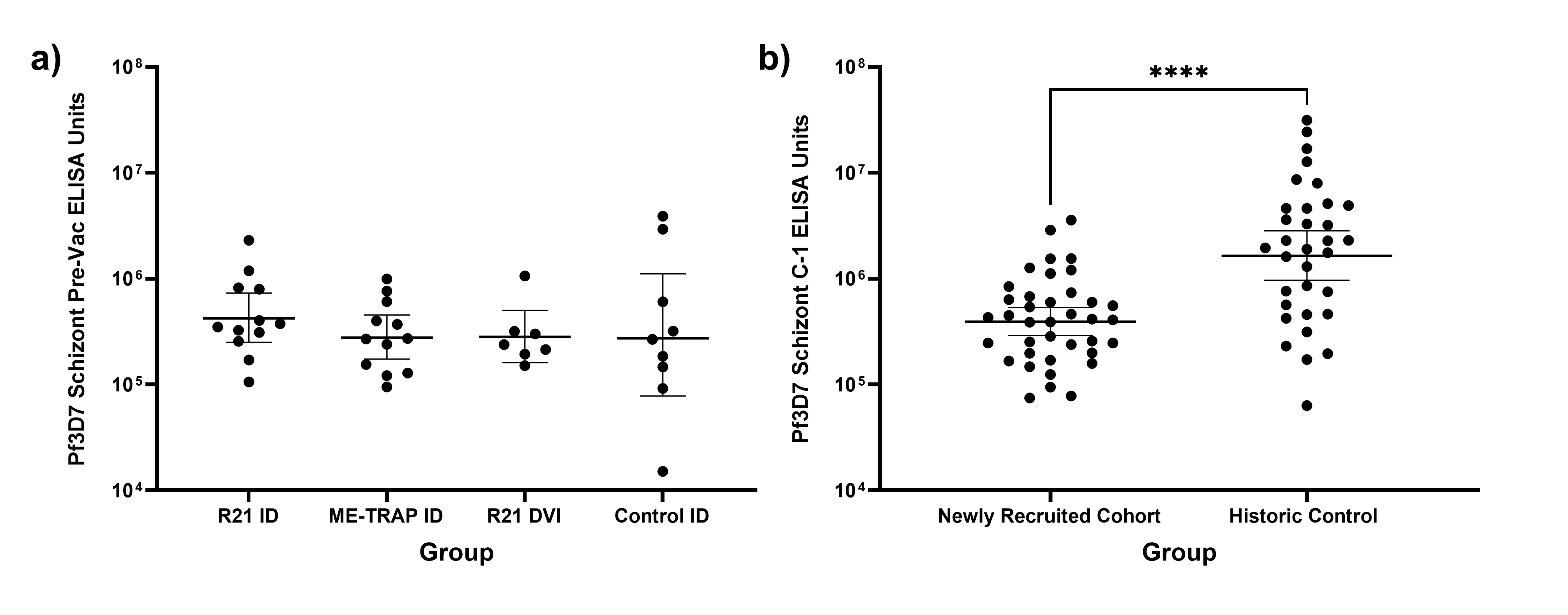

Schizont IgG ELISA units are shown measured at (a) pre-vaccination (Pre-Vac) and (b) day before challenge (C-1) against Pf3D7 schizont extract giving the geometric mean and error bars showing 95% confidence intervals.

#### Figure S5. Parasite growth curves following ID and DVI inoculation.

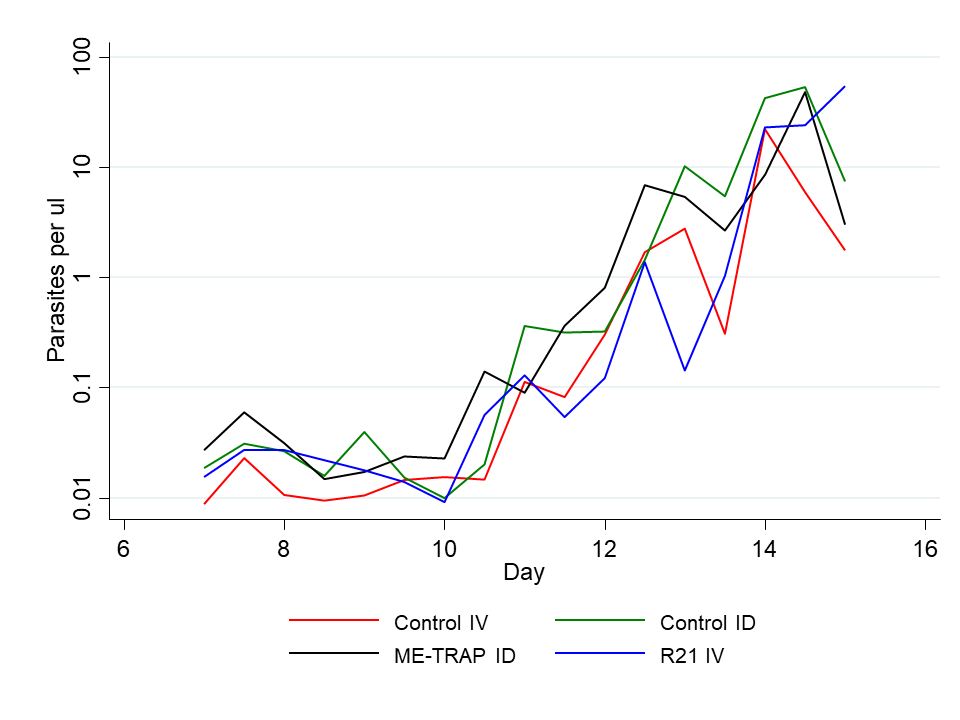

Geometric means of log PCR parasite densities following CHMI by group and day being R21 DVI (red line); Control ID (green line); ID ChAd6/MVA ME-TRAP (black line); and Historic Control DVI (blue line).
